# Cortical Thickness Patterns of Cognitive Impairment Phenotypes in Drug-Resistant Temporal Lobe Epilepsy

**DOI:** 10.1101/2023.08.04.23293483

**Authors:** Gadi Miron, Paul Manuel Müller, Louisa Hohmann, Frank Oltmanns, Martin Holtkamp, Christian Meisel, Claudia Chien

**Affiliations:** Epilepsy-Center Berlin-Brandenburg, Institute for Diagnostics of Epilepsy, Berlin, Germany; Epilepsy-Center Berlin-Brandenburg, Department of Neurology, Charité – Universitätsmedizin Berlin, Berlin, Germany; Computational Neurology, Department of Neurology, Charité – Universitätsmedizin Berlin, Berlin, Germany; Berlin Institute of Health, Berlin, Germany; Bernstein Center for Computational Neuroscience, Berlin, Germany; Center for Stroke Research Berlin, Berlin, Germany; NeuroCure Cluster of Excellence, Charité – Universitätsmedizin Berlin, Berlin, Germany; Experimental Clinical and Research Center, Charité – Universitätsmedizin Berlin, Berlin, Germany; Neuroscience Clinical Research Center, Charité – Universitätsmedizin Berlin, Berlin, Germany; Department of Psychiatry and Neuroscience, Charité – Universitätsmedizin Berlin, Berlin, Germany

## Abstract

**Background:** In temporal lobe epilepsy (TLE), a taxonomy classifying patients into three cognitive phenotypes has been adopted: minimally, focally, or generally cognitively impaired (CI). We examined grey matter (GM) thickness patterns of cognitive phenotypes in drug-resistant TLE and assessed potential use for predicting post-surgical cognitive outcomes.

**Methods:** TLE patients undergoing presurgical evaluation were categorized into cognitive phenotypes. Network edge weights and distances were calculated using ANOVA-III F-statistics from comparisons of GM regions between each TLE cognitive phenotype and age- and sex-matched healthy participants. In resected patients, logistic regression models (LRMs) based on network analysis results were used for prediction of post-surgical cognitive outcome.

**Results:** A total of 124 patients (63 females, mean age±SD=36.0±12.0 years) and 117 healthy controls (63 females, mean age±SD=36.1±12.0 years) were analyzed. In the generalized CI group (n=66, 53.2%), 28 GM regions were significantly thinner compared to HCs. Focally impaired patients (n=37, 29.8%) showed 13 regions, while minimally impaired patients (n=21, 16.9%) had 2 significantly thinner GM regions. Regions affected in both generalized and focally impaired patients included the anterior cingulate cortex, medial prefrontal cortex, medial temporal, and lateral temporal regions. In 69 (35 females, mean age±SD=33.6±18.0) patients that underwent surgery, LRMs based on network-identified GM regions predicted post-surgical verbal memory worsening with a receiver operating curve-area under the curve of 0.70±0.15.

**Conclusions:** A differential pattern of GM thickness can be found across different cognitive phenotypes in TLE. Including MRI with clinical measures associated with cognitive profiles has potential in predicting post-surgical cognitive outcomes in drug-resistant TLE.

## INTRODUCTION

In drug-resistant temporal lobe epilepsy (TLE), cognitive impairment (CI) is a major comorbidity that negatively affects the quality of life in 80% of patients^1, 2^. Traditionally, CI patterns in epilepsy were assumed to follow specific epilepsy syndromes and were studied in relation to clinical factors (e.g., duration of disease or the presence of hippocampal sclerosis). However, research has shown that cognitive dysfunction extends beyond clinical boundaries or syndrome-specific expectations, which provide only limited insight into patients’ cognitive profiles and prognoses^3^. To elucidate cognitive heterogeneity in epilepsy, standardize large-scale cognitive research efforts, and individualize treatment approaches, multiple studies have emphasized the importance of identifying unique patterns of CI, or cognitive phenotypes. This work has most recently culminated in the International Classification of Cognitive Disorders in Epilepsy (IC-CoDE) model^4^. IC-CoDE provides a robust framework that harmonizes cognitive diagnostics and classifies patients into three distinct cognitive phenotypes: minimal CI, focal impairment, and generalized multi-domain cognitive dysfunction^5–8^.

Large-scale neuroimaging studies in epilepsy have identified widespread patterns of brain atrophy compared to healthy controls (HCs)^9, 10^. However, these studies have not investigated specific structural differences related to different cognitive phenotypes. Prior studies with smaller cohorts have reported distinct patterns of cortical and subcortical changes in patients with a generalized impairment profile, whereas minimally impaired patients typically have minimal to no differences from HCs^11–13^. Furthermore, studies exploring cognitive phenotypes from a network topology perspective have indicated that impaired cognition may be related to increased disruption of cerebral network properties^3, 14, 15^. However, the relevance of these findings in clinical practice and decision-making is unclear. Particularly in patients undergoing presurgical assessment, an important question pertains to post-surgical cognitive prognosis and how this may be predicted. This has yet to be investigated within the framework of cognitive phenotyping and utilizing magnetic resonance imaging (MRI) derived cortical GM thickness measures.

Here, we (1) validate the cognitive phenotyping (IC-CoDE) approach in a large cohort of German patients with drug-resistant TLE, (2) identify differential GM thickness patterns per cognitive phenotypes, and (3) perform an exploratory analysis to investigate the utility of these clinical and MRI patterns for predicting post-surgical cognitive outcome.

## MATERIALS AND METHODS

### Participants

We conducted a retrospective analysis of patients that underwent presurgical evaluation between January 2012 and August 2022 at the Epilepsy-Center Berlin-Brandenburg (Figure 1). Patients were included in the study if they had drug-resistant TLE, had undergone neuropsychological assessment, and had an MRI following a uniform epilepsy imaging protocol. All included patients underwent anterior temporal lobectomy including resection of the hippocampus.

**Figure 1:**
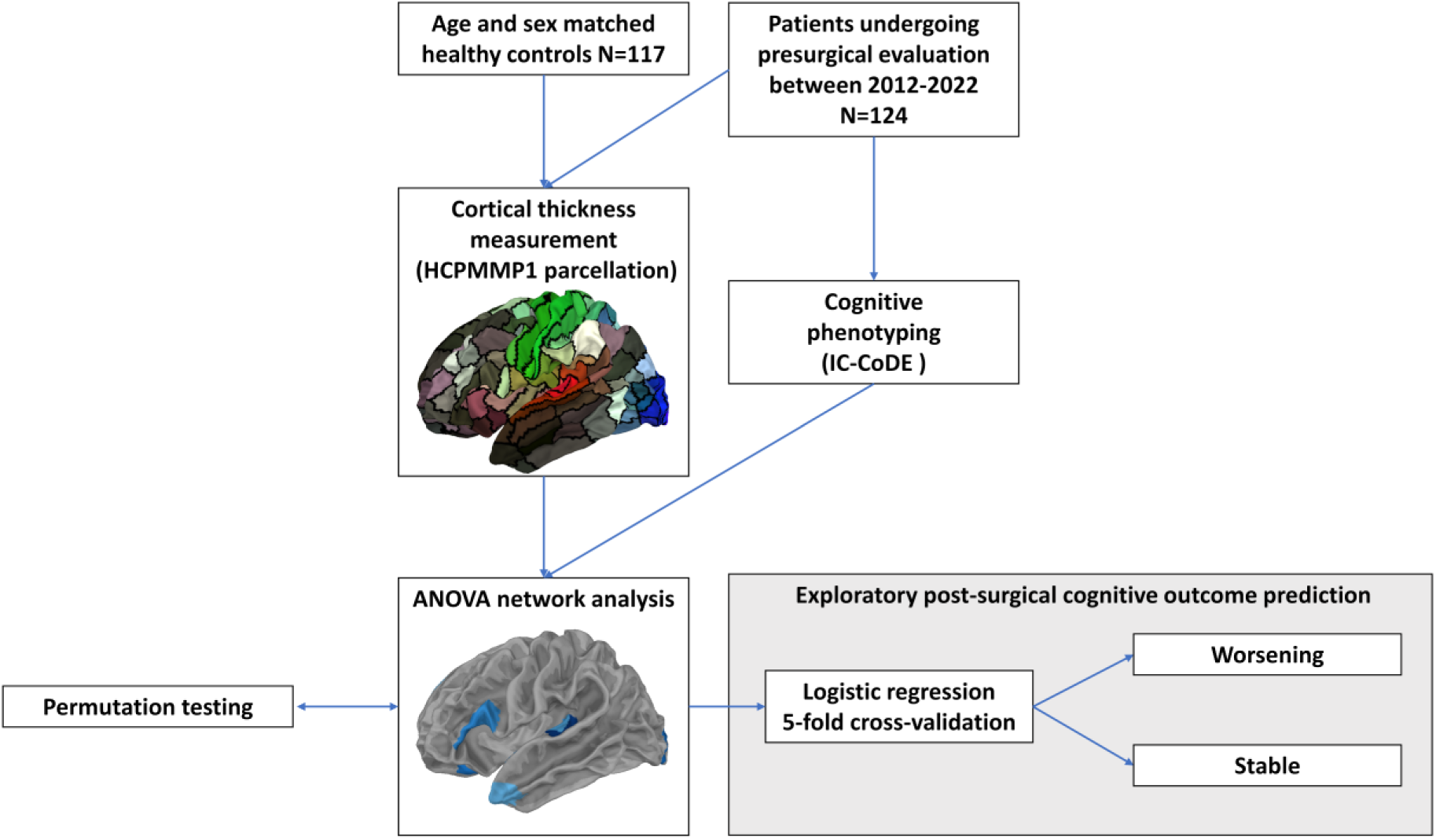
Study design. Cortical gray matter thickness measures were compared between patients with drug-resistant temporal lobe epilepsy and age- and sex-matched healthy controls accessed from the Human Connectome Project. A network analysis based on ANOVA comparison was performed to determine gray matter regions that were significantly different between patients and controls. These regions were then used to construct logistic regression predictors for post-surgical cognitive outcome. An additional permutation testing procedure was done to ensure validity of findings.

Controls were healthy subjects with imaging data downloaded from the publicly available Human Connectome Project (HCP) young adult, development and lifespan studies^18, 19^. Healthy controls (HC) were age- and sex-matched with the TLE cohort.

### Neuropsychological assessment

A total of 16 measures derived from routine neuropsychological testing were applied across five cognitive domains. Verbal learning and memory was evaluated using a word list learning test, the German version of the Rey Auditory Verbal Learning Test (“Verbaler Learn-und Merkfähigkeitstest”)^20^. Parameters assessed included immediate memory (VLMT1), reproduction of the last learning trial (VLMT5), total learning (sum of VLMT1-5) as well as delayed recall (VLMT7). Visual learning was examined using the Diagnostics for Cerebral Brain Injuries (“Diagnosticum für Cerebralschädigung”) and Recurring Figures Test^21, 22^, using the total of correctly reproduced patterns or figures, respectively, across all learning trials. Language was evaluated using semantic and phonetic word fluency tests with and without category switching (“Regensburger Wortflüssigkeitstest”)^23^. Working/short term memory was assessed by two block tapping tests and Mottier tests; and attention was assessed using the computerized Test battery for Attentional Performance (TAP; “Testbatterie für Aufmerksamkeitsprüfung”) with the subtests Alertness and Go/No-Go^24–26^. All test scores were normalized regarding age, sex, and/or education according to established norms provided in the test manuals and converted into percentile ranks. Patients that underwent surgery performed the same neuropsychological assessments one year after surgery.

### Cognitive phenotyping

Cognitive phenotyping according to the IC-CoDE criteria approach was applied to the TLE patient cohort prior to surgery^6^. Cognitive domain impairment was defined as two parameters within a domain having a score of at least one standard deviation (SD) below the mean of the norm population. Next, three cognitive phenotypes were established: (a) minimally impaired (MI) – no cognitive domain impaired; (b) focal impairment (FI) – one cognitive domain impaired; (c) generalized impairment (GI) – two or more cognitive domains impaired.

### MRI acquisition

All patients underwent 3-Tesla structural MRI scanning using a Magnetom Skyra (Siemens, Erlangen, Germany) scanner at Charité – Universitätsmedizin Berlin, Campus Benjamin Franklin. A standardized MRI protocol was performed, including a cerebral 3D T1-weighted sequences (TR/TE = 1900 ms/2.41 ms, TI = 900 ms, flip angle = 9°, voxel size = 0.875/0.875 mm). HC 3D T1-weighted cerebral MRIs were obtained from the Human Connectome Project (HCP) S1200 young adult release, HCP aging and development cohort with MRI sequence parameters as described in published imaging protocols^19, 27^.

### MRI data preprocessing

Epilepsy patient cerebral T1-weighted MRIs were converted to NIFTI format using dcm2niix, registered to MNI-152 space using fslreorient2std, N4-bias corrected using ANTs, and cropped using FSL robustfov (https://fsl.fmrib.ox.ac.uk/fsl/fslwiki/Fslutils)28–30. Since HCP data were already in NIFTI format, defaced and minimally processed, it should be noted that the MRIs were still preprocessed using the tools and cropped as the epilepsy patient T1-weighted MRIs. Cortical thicknesses were then parcellated using the CAT12 toolbox and subregional thicknesses were extracted using the HCP Multi-Modal Parcellation atlas, giving a total of 360 cortical regional thicknesses and volumes of the total intracranial region, total intracranial GM, and total intracranial white matter^18, 31^.

### Statistical analysis

#### ANOVA-based cortical thickness analysis

Type-III ANOVA models were used to compare patients from each cognitive phenotype group to a matched HCs subgroup, with MRI regions of interest (ROIs) measures as response variables. Age and sex were included as covariates as well as total intracranial volume and MRI scanner to address technical confounders of cortical thickness measures. For all analyses, p-values were Bonferroni-corrected for multiple comparison by the total number of ROIs (n=363). Clinical covariates assessed within patients in each cognitive phenotype also included: epilepsy duration (years), total lifetime number of anti-seizure medications (ASMs), history of focal to bilateral tonic-clonic seizures (FBTCS), and seizure onset side. To assess the robustness of our results, we conducted a permutation test in addition to the standard ANOVA analysis (Supplementary Methods).

Next, we examined ANOVA results using a directed multimodal network analysis approach^33^. Directed links were constructed from clinical (including cognitive phenotype group) and confounding measures to MRI ROIs and edge weights were the F-statistics obtained from the (non-permuted) ANOVA type-III models. Only nodes with significant connections (edge weights falling within a 95% F-distribution) were used to build directed network graphs for each cognitive cluster. Distances between nodes were defined per cognitive phenotype as average edge weight divided by the individual edge weight, with shorter distances reflecting a more robust connection difference between TLE patient and HC. To account for possible confounders related to heterogeneous MRI collection, we excluded ROIs closely connected to MRI (distances below 2).

#### Exploratory examination of post-surgical cognitive outcome prediction

We conducted an additional analysis for 1-year post-surgical cognitive outcome prediction for each cognitive domain. Cognitive worsening was defined as a decrease in post-surgical percentile of more than 5% compared to the pre-surgical score. For predictions, the independent variables were the combined ROIs (n=13) identified as most significantly connected to each cognitive phenotype (distances below 1), as post-surgical change was similarly distributed across cognitive profiles and splitting into separate phenotypic groups would yield sample sizes that are insufficient for predictive modelling. First, descriptive LRMs were fitted to cognitive outcomes in each domain. Second, to determine if these regions along with clinical parameters could be predictive of cognitive changes, we applied 100 times repeated 5-fold nested cross-validation with recursive feature elimination and l2 regularizations. To assess model performance, we compared them to two random classifiers. The first established chance level by randomly assigning predictions classes to patients while maintaining the label prevalence of the data. The second classifier utilized random ROIs instead of the original factors selected as features (keeping n=13). Different LRMs were compared using the receiver operating curve-area under the curve (ROC-AUC) values calculated from the hold-out dataset predictions of the cross-validation scheme (see additional details in Supplementary Methods).

### Code availability

Analysis code is available at https://gitlab.com/computational-neurologie/structural_cognitive_TLE.

### Data availability statement

All HC data was downloaded from the freely available HCP dataset (https://www.humanconnectome.org/). Patient data are not publicly available due to privacy restrictions but may be made available upon reasonable request to the corresponding author.

### Ethics approval

This study was approved by the Institutional Review Board of Charité – Universitätsmedizin Berlin (reference number EA2_084_22). Due to the retrospective nature of the study, informed consent of patients was waived.

## RESULTS

### Participants

One hundred and twenty-four patients (63 females, mean age 36.0±12.0 years) and 117 age- and sex-matched healthy controls (63 females, 36.1±12.0 years) were included in the study. Patient demographics and clinical characteristics are shown in Table 1.

**Table 1.** Baseline patient demographics

|  | All patients | Generalized impaired | Focally impaired | Minimally impaired | P- value |
| --- | --- | --- | --- | --- | --- |
| Number of patients | 124 | 66 | 37 | 21 |  |
| Sex F (M) | 63(61) | 30(36) | 19(18) | 14(7) | 0.2 |
| Age | 36.0±12.0 | 34.6±11.6 | 39.4±12.1 | 34.3±12.3 | 0.1 |
| Epilepsy duration (years) | 18.0±13.2 | 18.4±14.0 | 18.4±13.2 | 14.2±10.5 | 0.8 |
| Education |  |  |  |  | 0.01* |
| Twelve years or less | 47 | 31 | 12 | 4 |  |
| Post-secondary non-tertiary education | 41 | 25 | 10 | 6 |  |
| Completion of secondary education | 12 | 3 | 4 | 5 |  |
| Academic education | 24 | 7 | 11 | 6 |  |
| History of FBTCS | 96 | 52(14) | 31(6) | 13(8) | 0.1 |
| Total number of ASMs (previous and current) | 5.0±2.2 | 5.3±2.2 | 5.1±2.1 | 4.2±1.6 | 0.2 |
| MRI findings |  |  |  |  | 0.5 |
| MTS | 46 | 28 | 11 | 7 |  |
| Non-lesional | 55 | 25 | 19 | 11 |  |
| Others (focal cortical dysplasia, low grade tumor, vascular lesions, lesions of unclear significance) | 23 | 13 | 7 | 3 |  |
| Seizure onset side |  |  |  |  | 0.6 |
| Left | 65 | 37 | 20 | 8 |  |
| Right | 49 | 25 | 14 | 10 |  |
| Bilateral | 10 | 4 | 3 | 3 |  |
| Resected, yes(no) | 77(47) | 45(21) | 18(19) | 14(7) | 0.1 |
| Surgical outcome |  |  |  |  | 0.8 |
| Engel 1 | 59 | 33 | 15 | 11 |  |
| Engel 2-4 | 17 | 12 | 2 | 3 |  |

Abbreviations: ASM: anti-seizure medications; FBTCS: focal to bilateral tonic-clonic seizures; MRI: magnetic resonance imaging; MTS: mesial temporal sclerosis. \*Significant differences ( $p < 0.05$ from an $\chi^2$ test).

### Cognitive clustering

Patients were classified into three groups: generalized impaired (GI, 66 patients 53.2%), focally impaired (FI, 37 patients, 29.8%) and minimally impaired (MI, 21 patients, 16.9%; Table 1 and Figure 2). Education level was different between the three phenotypes, with GI patients having achieved a significantly lower education level than both the FI and MI patients (p = .01, Table 1).

**Figure 2:**
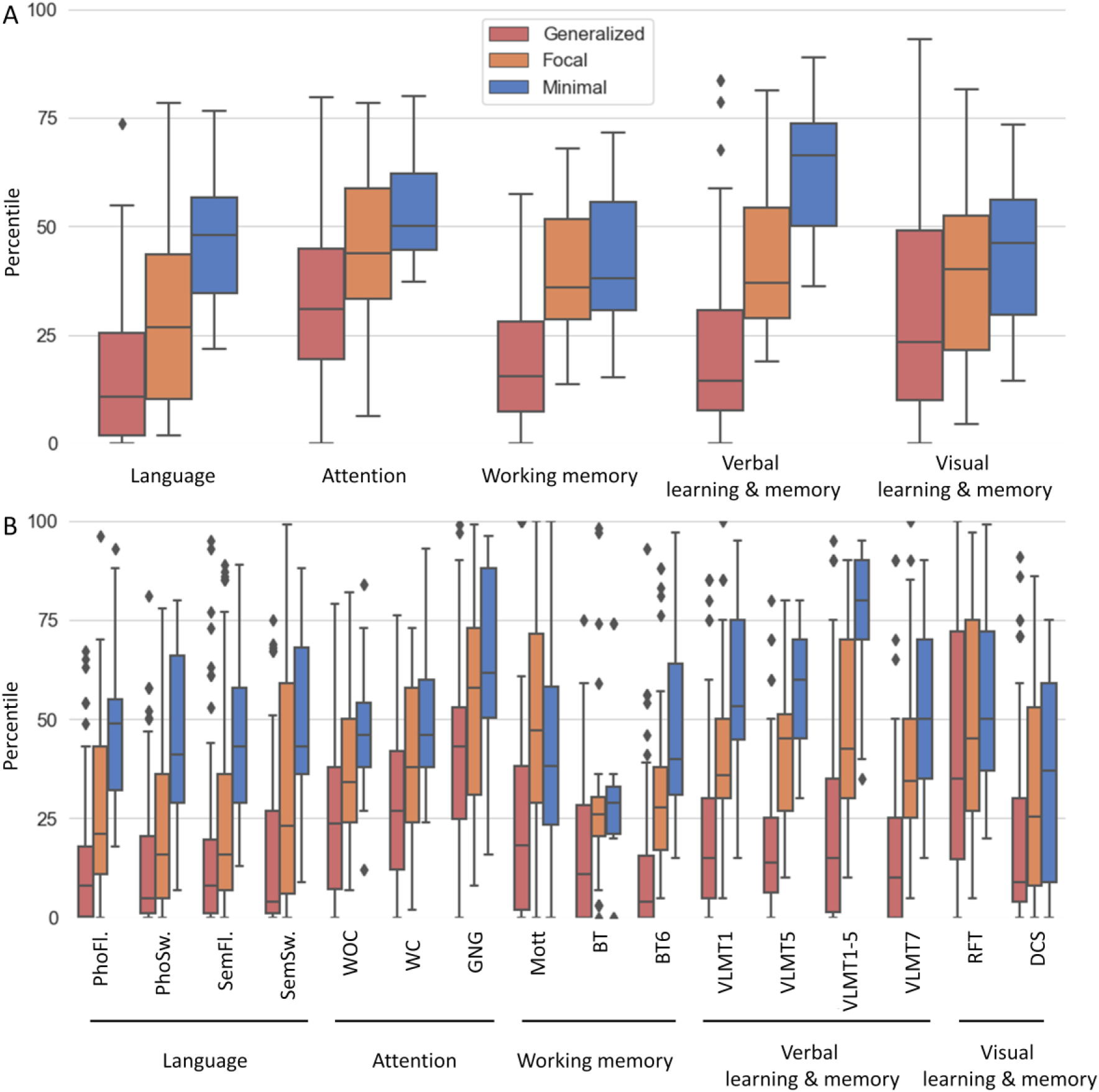
Cognitive phenotyping. A. Mean percentile scores for each domain and cognitive phenotype. B. Means percentile scores for each cognitive test and cognitive phenotype. Y axis shows percentile ranks in comparison to healthy norm populations. Boxes show median, interquartile range, whiskers show 1.5 times the interquartile range. Generalized impairment group shown in red, focal impaired shown in orange, and minimal impaired in blue. Abbreviations: tests per domain: language: PhoFl. – phonetic word fluency, PhoSw. - phonetic category switching, SemFl. – semantic word fluency, SemSw. - semantic category switching; attention: WOC- Testbatterie für Aufmerksamkeitsprüfung (TAP) Alertness subtest without auditory warning cue, WC- with auditory warning cue, GNG-Go/No-Go subtest; verbal learning and memory: VLMT - Verbaler Lern- und Merkfähigkeitstest; visual learning and memory: DCS-Diagnosticum für Cerebralschädigung test, RFT-Recurring Figures Test; Working/short term memory-BT-Block Tapping, Mott-Mottier test.

### Cortical thickness patterns associated with cognitive phenotypes

Each cognitive group was compared to age- and sex-matched HCs (complete results of ANOVA analyses are provided in Supplementary Table 1). ROIs and corresponding distances from network analysis are presented in Figure 3 and Supplementary Table 2.

**Figure 3:**
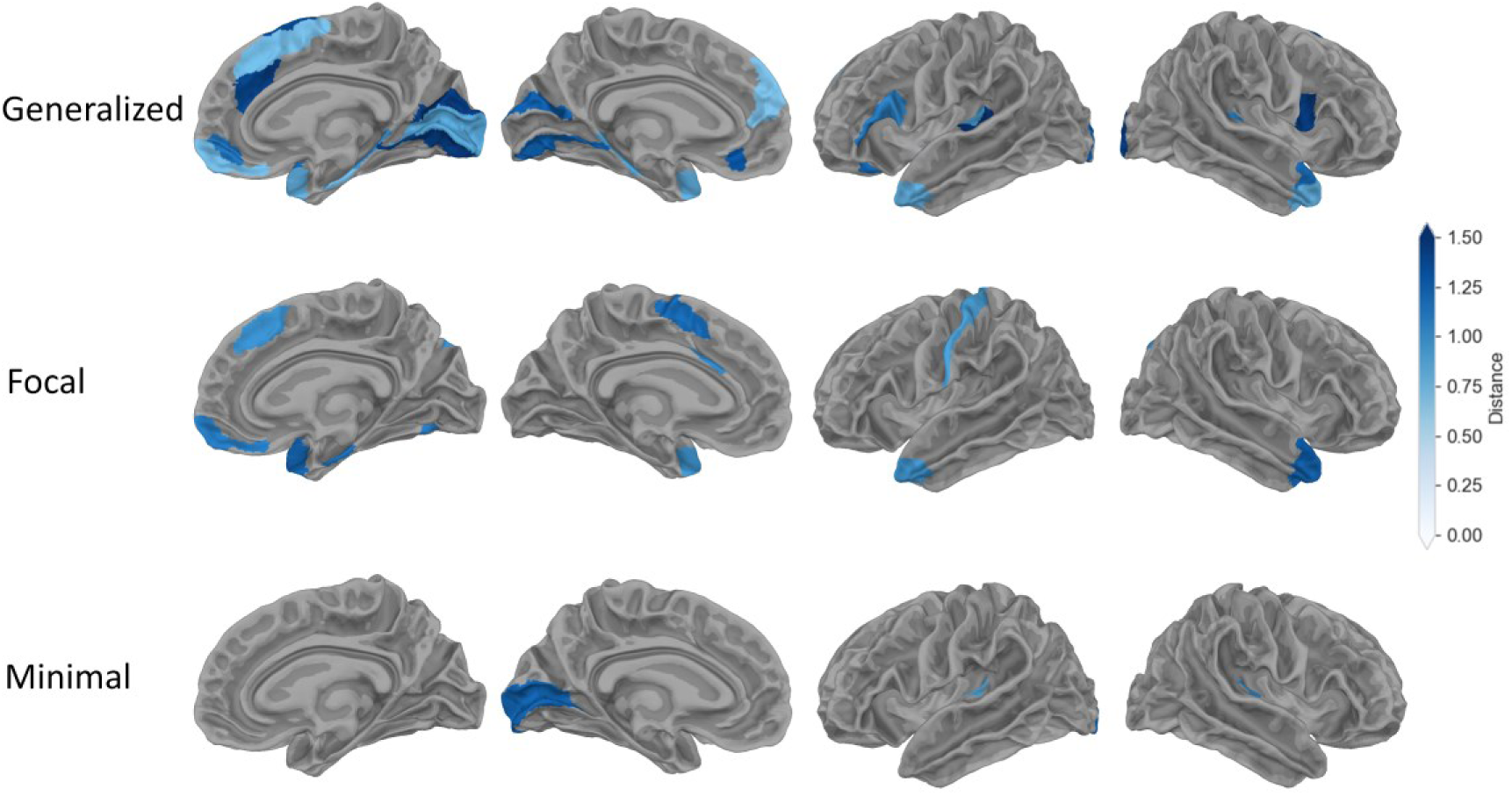
Cortical thickness abnormalities in drug-resistant TLE patients. The plots show cortical grey matter regions with significant differences from age- and sex-matched healthy controls for the three cognitive phenotypes. Distance is defined as the average F-statistic / F-statistic, with shorter distances reflecting a more robust difference between cognitive impaired patient and healthy controls. Shorter distances are reflected by lighter shades of blue.

In the GI group, 28 ROIs were significantly affected, primarily within the bilateral anterior cingulate and medial prefrontal cortex (7 ROIs), bilateral early auditory cortex (4 ROIs), bilateral medial (3 ROIs) and lateral temporal regions (2 ROIs), bilateral auditory association (2 ROIs), left inferior frontal (2 ROIs) and bilateral early visual (2 ROIs).

The FI group showed 12 affected ROIs, overlapping with the GI group by 7 ROIs. These included ROIs located in the bilateral anterior cingulate and medial prefrontal (3 ROIs), right medial (1 ROI) and bilateral lateral temporal regions (2 ROIs), bilateral auditory association (2 ROIs), left paracentral lobular and mid cingulate (1 ROI), left somatosensory and motor (1 ROI), right ventral stream visual (1 ROI), right dorsal stream visual (1 ROI).

For the MI group, only 3 ROIs were affected in the bilateral early auditory (2 ROIs) and left primary visual left (1 ROI) cortical regions, where the left early auditory ROI was also found in the GI patient group.

Cortical thickness was decreased in CI groups compared to HCs in all significantly affected regions except the left somatosensory and motor cortex in the FI group (see effect sizes in Supplementary Figure 1). Examining cortical thickness measures affected by CI with clinical and demographic variables, we found that age was also connected to 7 ROIs (Supplementary Table 1, complete results for age in Supplementary Figure 2). The remaining clinical covariates were not significantly connected to cortical GM thickness.

To assess the robustness of these findings, permutation testing was performed prior to re-running our ANOVA-based network analysis in each cognitive cluster. Shuffling the ROI labels from each patient removed any significant association of the ROIs, indicating that the initial, true analysis with original labels and the observed results were unlikely to have arisen merely by chance.

### Post-surgical cognitive outcomes

Sixty-nine patients underwent surgery and post-surgical cognitive testing (Table 2, Figure 4). Across all cognitive domains, lower performance in presurgical cognitive testing was associated with improved post-surgical cognitive performance (Table 2).

**Table 2.** Post-surgical demographics and cognitive outcomes

Abbreviations: ASM: anti-seizure medications; FBTCS: focal to bilateral tonic-clonic seizures;
|  | Total | Language (N=68) |  | Working memory (N=66) |  | Verbal memory (N=64) |  | Non verbal memory (N=68) |  |
| --- | --- | --- | --- | --- | --- | --- | --- | --- | --- |
|  |  | Non worsening (N=51) | Worsening (N=17) | Non worsening (N=54) | Worsening (N=12) | Non worsening (N=40) | Worsening (N=24) | Non worsening (N=49) | Worsening (N=19) |
| Sex, female(male) | 35(34) | 27(24) | 7(10) | 27(27) | 6(6) | 20(20) | 12(12) | 24(25) | 10(9) |
| Age | 33.6 ± 18.0 | 32.6±11.8 | 35.2±11.4 | 32.6±10.9 | 31.2±9.5 | 32.8±9.9 | 29.9±10.0 | 33.1±11.8 | 33.6±11.5 |
| Epilepsy duration | 18.0 ± 14.1 | 17.8±14.2 | 16.5±11.4 | 18.4±13.7 | 10.8±8.4 | 17.6±12.6 | 14.2±12.1 | 18.5±14.3 | 14.8±10.7 |
| Total number of ASMs | 4.9±2.3 | 5.2±2.5* | 3.9±1.6 | 4.8±2.3 | 4.9±2.0 | 4.8±2.5 | 4.8±1.8 | 5.1±2.4 | 4.5±2.1 |
| History of FBTCS, yes (no) | 51(18) | 14(37) | 4(13) | 14(40) | 2(10) | 12(28) | 4(20) | 13(36) | 5(14) |
| Pre-surgical domain percentile |  | 18.5±18.3**<br>* | 40.2±21.1**<br>* | 23.0±15.6**<br>* | 41.8±19.1**<br>* | 27.4±23.6** | 41.9±24.5** | 30.6±22.7**<br>* | 53.3±19.6**<br>* |
| Cognitive phenotype |  |  |  |  |  |  |  |  |  |
| Generalized impaired | 38 | 31 | 6 | 33 | 4 | 23 | 13 | 26 | 11 |
| Focally impaired | 21 | 11 | 7 | 11 | 6 | 11 | 5 | 13 | 5 |
| Minimally impaired | 10 | 9 | 4 | 10 | 2 | 6 | 6 | 10 | 3 |
| Resection side, left(right) |  | 23(28) | 10(7) | 26(28) | 8(4) | 16(24) | 16(8) | 25(24) | 8(11) |
| Surgical outcome, Engel 1(Engel 2-4) | 56(13) | 43(8) | 12(5) | 43(11) | 10(2) | 32(8) | 19(5) | 41(8) | 14(5) |
| MTS pathology (non MTS) |  | 20(31) | 3(14) | 18(36) | 5(7) | 13(27) | 8(16) | 20(29) | 3(16) |

MTS: mesial temporal sclerosis. Statistical differences were assessed with regard to worsening or non-worsening within each cognitive domain \*p < 0.05, \*\*p<0.01, \*\*\*p<0.001, Mann-Whitney U test.

**Figure 4:**
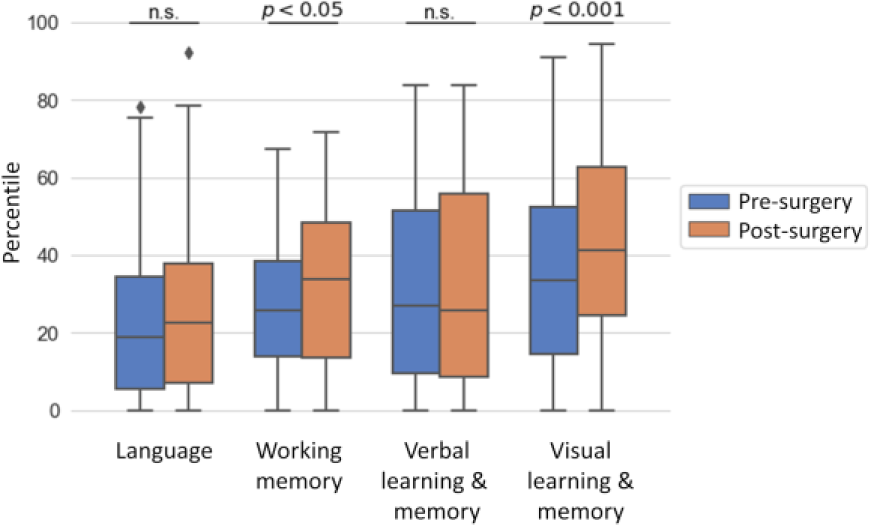
Cognitive changes after epilepsy surgery. Sixty-nine patients underwent surgery. Orange boxes display pre-surgical and blue boxes show post-surgical percentiles per domain.

### Exploratory analysis of cortical thickness patterns with 1-year post-surgical cognitive worsening

Having identified ROIs related to each cognitive phenotype group, and since the same type of epilepsy surgery was performed on all patients, we applied multivariable logistic regression analysis to assess if cortical thickness measures could confer information about the risk of post-surgical cognitive worsening (Figure 5). The ROIs we included in this analysis are presented in Table 3, as well as significant predictors in the verbal memory cognitive domain. Results for the remaining cognitive domains are presented in Supplementary Figure 3.

**Figure 5:**
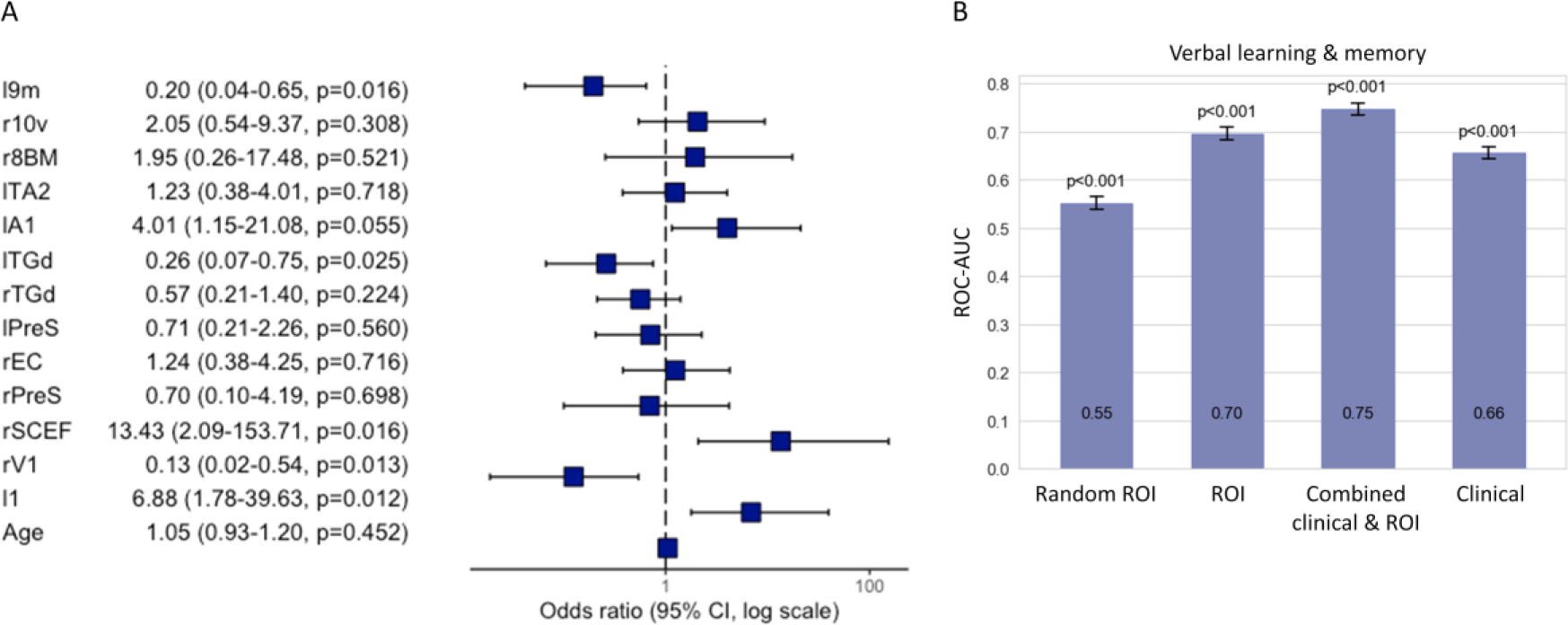
Descriptive and predictive logistic regression models of verbal memory post-surgical outcome. A. Odds ratios for descriptive logistic regression classifying the change of verbal memory after surgery using the regions strongest associated with cognitive phenotype and age as independent variables. The cortical thicknesses are rescaled with respect to the mean and standard deviation of the healthy controls. B. Area under the receiver operating curve (ROC-AUC) of predictive logistic regression models with recursive feature reduction and l2 regularization for changes of verbal memory after surgery evaluated through repeated, nested cross-validation. Models displayed include: random ROI-predictor based on randomly chosen brain regions; ROI-predictor based on cognitive phenotype significantly associated regions; clinical-predictor based on clinical variables; combined clinical and ROI. Models were significantly different from a chance level classifier and among each other (p < 0.001).

**Table 3.** Cortical grey matter regional thicknesses evaluated for post-surgical cognitive prediction

| Area name | Area description | Side | Lobe | Cognitive phenotype | Cognitive cluster distance | Odds ratio - verbal memory outcome prediction |
| --- | --- | --- | --- | --- | --- | --- |
| 9m | Anterior cingulate and medial prefrontal | Left | Frontal | Generalized | 0.72 | 0.2 (0.04-0.65, p=0.02)* |
| 1 | Somatosensory and motor | Left | Parietal | Focal | 0.92 | 6.88 (1.78-39.6, p=0.01)* |
| TA2 | Auditory association | Left | Temporal | Focal | 0.99 | 1.23 (0.38-4.01, p=0.7) |
| A1 | Early auditory | Left | Temporal | Generalized | 0.92 | 4.01 (1.15-21.08, p=.06) |
| TGd | Lateral temporal | Left | Temporal | Focal | 0.97 | 0.26 (0.07-0.75, p=0.03)* |
| PreS | Medial temporal | Left | Temporal | Generalized | 0.89 | 0.71 (0.21-2.26, p=0.6) |
| 10v | Anterior cingulate and medial prefrontal | Right | Frontal | Generalized | 0.74 | 2.05 (0.54-9.37, p=0.3) |
| 8BM | Anterior cingulate and medial prefrontal | Right | Frontal | Focal | 0.75 | 1.95 (0.26-17.48, p=0.5) |
| SCEF | Paracentral lobular and mid cingulate | Right | Frontal | Generalized | 0.73 | 13.43 (2.09-153.71, p=0.02)* |
| V1 | Primary visual | Right | Occipital | Generalized | 0.86 | 0.13 (0.12-0.54,<br>p=0.01)* |
| TGd | Lateral temporal | Right | Temporal | Generalized | 0.82 | 0.57 (0.21-1.4,<br>p=0.2) |
| EC | Medial temporal | Right | Temporal | Generalized | 0.74 | 1.24 (0.38-4.25,<br>p=0.7) |
| PreS | Medial temporal | Right | Temporal | Generalized | 0.98 | 0.7 (0.21-2.26,<br>p=0.6) |

\*Odds ratio is significant with $p < .05$

### Exploratory predictive performance of post-surgical cognitive outcomes

Finally, we evaluated the predictive performance of a LRM using a 5-fold nested cross-validation (with 100 repetitions) scheme. Specifically, using the 13 identified cortical regions as features (Table 3), we found that post-surgical verbal memory could be predicted with an ROC-AUC of 0.70±0.15 and an accuracy of 0.65±0.13. Notably, the MRI-extracted cortical thickness-based prediction model outperformed that which was based on clinical parameters alone (ROC-AUC 0.62±0.14), as well as models based on random ROIs (Figure 5B). Combining clinical features with MRI measures further improved predictive performances to ROC-AUC 0.75±0.14 and accuracy 0.69±0.12. To rule out that the results are an artefact of the chosen threshold for worsening, we examined additional different thresholds, which remained well above chance level, see Supplementary Figure 5. Prediction models used to test cognitive outcomes related to visual memory, working memory, and language performed no better than chance.

## DISCUSSION

We here identified cognitive phenotypes in a large German cohort of patients with drug-resistant TLE. Through a graph theory-based analysis of MRI and clinical measures, we found different grey matter thickness patterns related to the degree of cognitive impairment. We examined 180 brain regions per hemisphere and delineated specific temporal and extratemporal cortical brain regions that may share relationships with CI, as well as those uniquely affected in each cognitive profile. Finally, we explored the potential use of cortical GM regions which we identified for clinically relevant prediction of post-surgical cognitive outcome in drug-resistant TLE patients.

Our study presents one of the first expertly curated German cohorts with cognitive phenotyping that underwent a broad battery of neuropsychological tests spanning five cognitive domains. Cognitive phenotyping has previously been established in a large American English speaking cohort^4^, and recently validated in an American Spanish speaking cohort^34^. Our study shows broader applicability of the IC-CoDE criteria approach and thus aids in the undertaking to standardize cognitive research across countries and languages. In our cohort, the largest group of patients was categorized into the GI (53.2%), followed by FI (29.8%) and finally MI (16.9%) cognitive groups. Compared to prior studies, we observed a higher percentage of GI patients, possibly due to the medication-resistant profile and long average epilepsy duration (18.0±13.2 years) of the entire cohort^7^. This population was specifically chosen to identify MRI markers that have clinical implications in presurgical evaluation. However, even within this highly resistant population, one in six patients had minimal CI, emphasizing the heterogeneity of cognitive function in TLE.

Our results point to distinct, severity-associated reduction of cortical thickness in specific GM regions with respect to CI. GI patients had the most cortical GM regions that differed from HCs, followed by FI and MI patients. Importantly, these patterns were independent of classical TLE-related clinical characteristics, underscoring pathophysiological differences between cognitive phenotype groups and emphasizing the benefit of a personalized cognitive approach in presurgical evaluation of TLE. There have also been observations in other MRI studies that reported increased diffusion and fMRI abnormalities in GI patients compared to MI patients^11, 15, 35^, as well as studies that reported a greater disturbance of functional network measures with increased impaired cognition^10, 14^. To date, relatively few studies have examined cortical thickness regional differences in cognitive phenotypes that involve adult drug-resistant TLE patients^3, 12, 35^. In two prior studies that examined a relatively benign TLE cohort, only modest associations with cognitive phenotypes were found, in contrast both to our findings and to an another study that specifically focused on a group of patients with highly medication-resistant TLE^3, 12^. These contrasting results highlight the importance of distinguishing between drug-resistant and responsive patients, and suggests that in impaired cognitive phenotypes (i.e., GI or FI), potential differences in structural abnormalities may exist between drug-responsive and refractory patients. Whereas MI patients have more similar MRI characteristics to HC regardless of drug-responsive status.

We identified 18 temporal and 25 extratemporal cortical brain regions affected in the impaired cognitive profiles. We observed an overlap of 7 ROIs between the FI and GI patient groups, indicating the existence of both shared and distinct structural cortical network changes that contribute to different severities of cognitive deficits. Both impaired phenotypes exhibited reduced cortical thickness of the lateral and medial temporal regions, known to be related to cognitive functions that include memory, learning and language^36^. Auditory cortical regions, were also shared between impaired phenotypes, which underscores the importance of auditory processing in contributing to difficulties in language and verbal memory, i.e., common cognitive domains affected in TLE^7^. Notably, the anterior cingulate and medial prefrontal cortical regions exhibited the most bilaterally widespread differences with HC, where 10 significant ROIs (7 in GI and 3 in FI) were identified. Previous studies have found that in patients with TLE these regions are associated with impaired memory, cognitive slowing, and attention ^36–38^. Adding to these studies, we demonstrated that these cortical regions are affected in CI of drug-resistant TLE, contextualising the role of these regions within the cognitive taxonomy framework. Although the MI group showed minimal cortical thickness alterations compared to HCs, it is notable that reduced cortical thickness was observed in the bilateral early auditory and primary visual cortices. These regions were also affected in the GI group (early auditory – 4 ROIS, early visual – 2 ROIS). Together, our findings raise the possibility that basic sensory processing may play a role in cognitive deficits in TLE. In the GI group, the distinguishing characteristics from other phenotypes were extensive involvement within each brain GM region (i.e., multiple ROIs identified as significantly different from HCs within several cortical brain region). Also, additional frontal lobe regions were found to be affected, including the orbital and polar frontal lobe, and dorsolateral prefrontal regions.

In patients with drug-resistant TLE undergoing presurgical evaluation, a crucial clinical task is to determine the risk for post-surgical cognitive worsening, which is the most common comorbidity associated with the procedure^39^. Utilising 13 cortical thickness measures, we were able to moderately predict 1-year post-surgical outcomes of verbal memory in TLE patients. Importantly, we found that combining clinical characteristics and imaging measures led to a significant improvement in the prediction performance, achieving an ROC-AUC of 0.75. Although we consider our analysis to be hypothesis-generating and exploratory, our findings suggest that multimodal models may offer additional value for pre-surgical cognitive risk assessments. Currently, state-of-the-art cognitive outcome prediction relies on subjective clinical assessment and neuropsychological testing alone^16^. Prior studies that related cognitive phenotypes to imaging abnormalities^41–43^, have yet to be applied for prediction of post-surgical verbal memory outcome. One study has used a latent disease factor-approach to perform a prediction of post-surgical inter-patient variability of verbal IQ. However, compared to this approach, our study requires only one (rather than four) imaging measures and provides a risk assessment of post-surgical worsening^17^. Notably, cognitive phenotypic groups were not associated with the post-surgical cognitive outcome, which is in-line with previous studies on patients with drug-resistant TLE^40^.

Our study has several limitations. First, we age- and sex-matched control data from the publicly available HCP which may not fully represent the local population from which our patient cohort was drawn. Secondly, MRI protocols and scanners were different in the healthy control cohort. However, to account for these differences we included MRI scanner as a variable in all our analyses, used a conservative multiple comparisons correction method during the ANOVA analysis as well as a permutation robustness-testing procedure. We observed relatively few significant MRI-extracted differences between the MI group and the controls, suggesting our findings are likely not due to data collection from different centers. Thirdly, cognitive tests used for phenotyping were limited by the availability of data. Thus, it was not possible to include a naming test in the language domain, and instead of the visuospatial domain used in the IC-CoDE criteria we used visual learning. However, cognitive phenotyping is aimed at bridging the gap caused by variability in tests between different countries and languages, and clear cognitive profiles were identified nevertheless. For cognitive outcome predictions, we defined worsening as an average deterioration of over five percentile points across tests in a domain. Bias may be introduced from averaging different measures within a domain, and we could not account for differences resulting in measurement error. However, using change in percentile ranks rather than raw scores, we took into account the patients’ age, sex and education, and we report consistent prediction results for additional thresholds. Lastly, a limitation arises due to the retrospective nature of this study, where clinical factors possibly affecting cognitive scores such as recent seizures, medication changes, sleep patterns, and mood could not be assessed.

## CONCLUSIONS

We found cognitive phenotypes in a large drug-resistant TLE cohort where links to differential patterns of cortical grey matter regional changes could be identified. In each cognitive profile, distinct cortical regions are significantly altered compared to age- and sex-matched HCs. We show that cortical thickness patterns and measures are associated with cognitive profiles and that there is clinical potential for use of these metrics for the prediction of post-surgical verbal memory.

### Contributorship statement

G.M, P.M, C.C, and C.M conceived and designed the study, contributed to data analysis and interpretation, wrote and critically revised the manuscript. M.H, L.H, F.O were involved in patient care, collection of data, and critically revising the manuscript. G.M collected the data, produced the original manuscript that all authors approved, and takes full responsibility for the overall content as the guarantor.

## Data Availability

All healthy control data was downloaded from the freely available HCP dataset (https://www.humanconnectome.org/). Patient data are not publicly available due to privacy restrictions but may be made available upon reasonable request to the corresponding author.

