## Supplementary figures and images for "Cortical Thickness Patterns of Cognitive Impairment Phenotypes in Drug-Resistant Temporal Lobe Epilepsy"

### Supplementary Figure 1

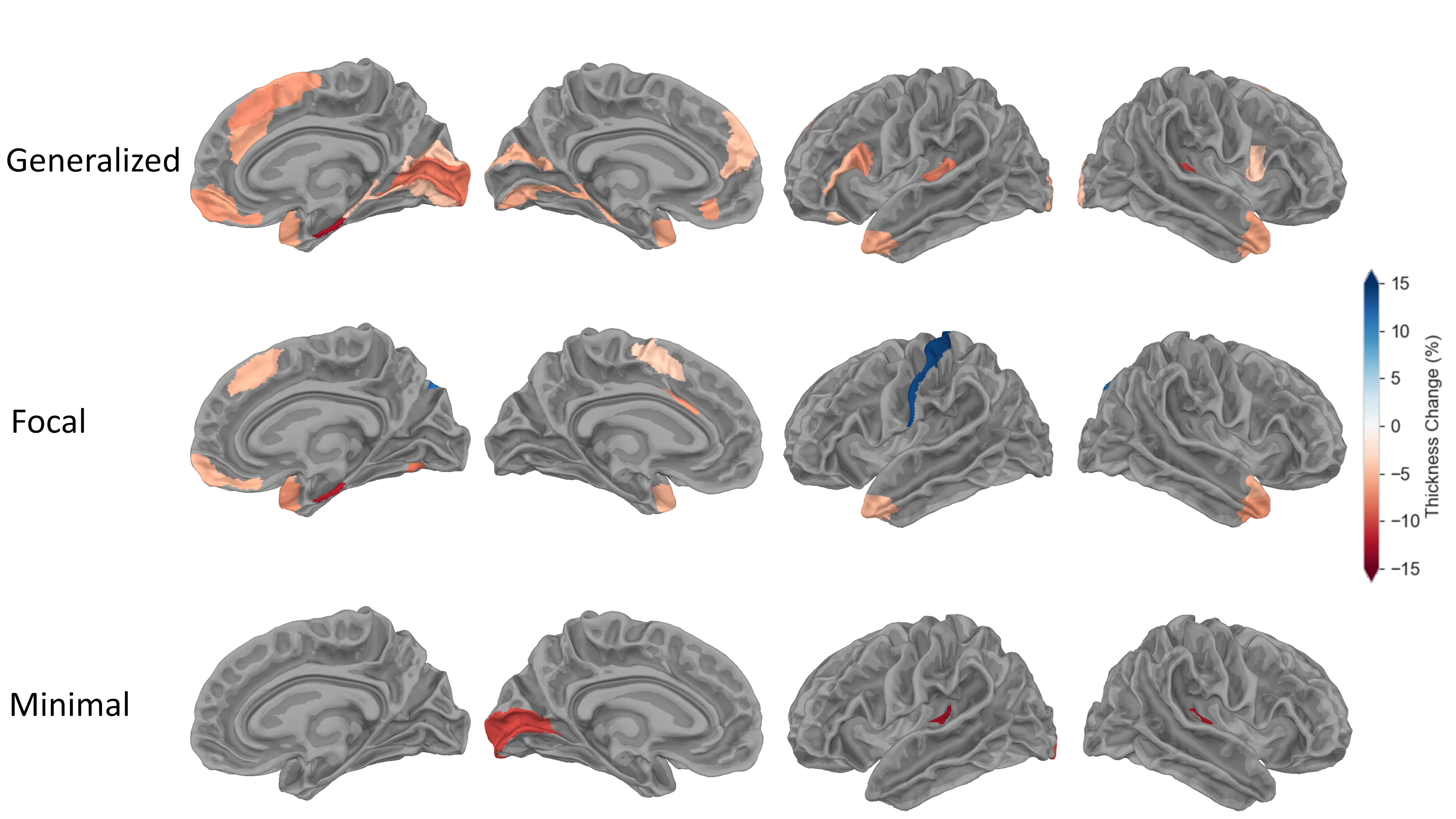

### Supplementary Figure 2

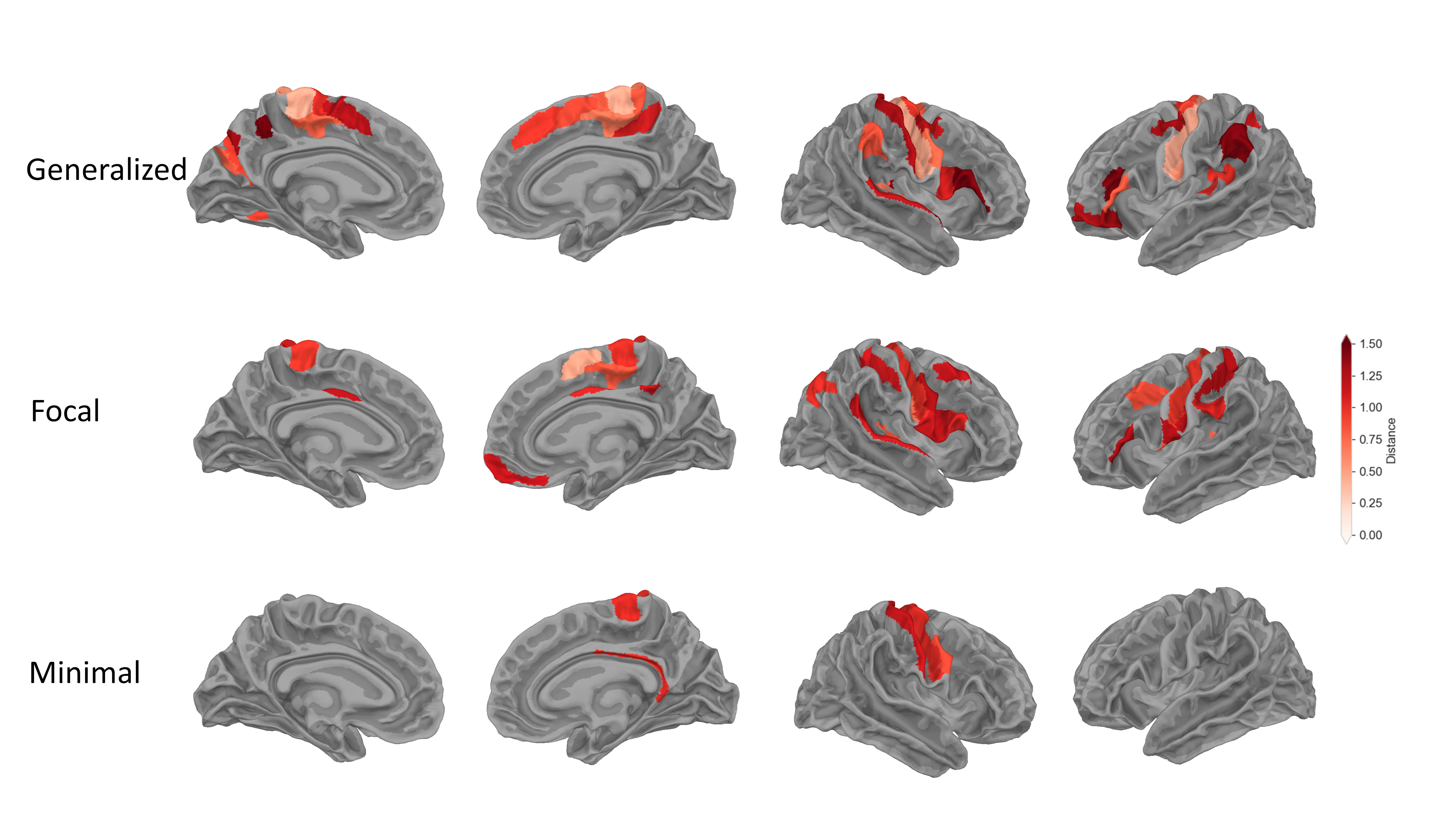

### Supplementary Figure 3

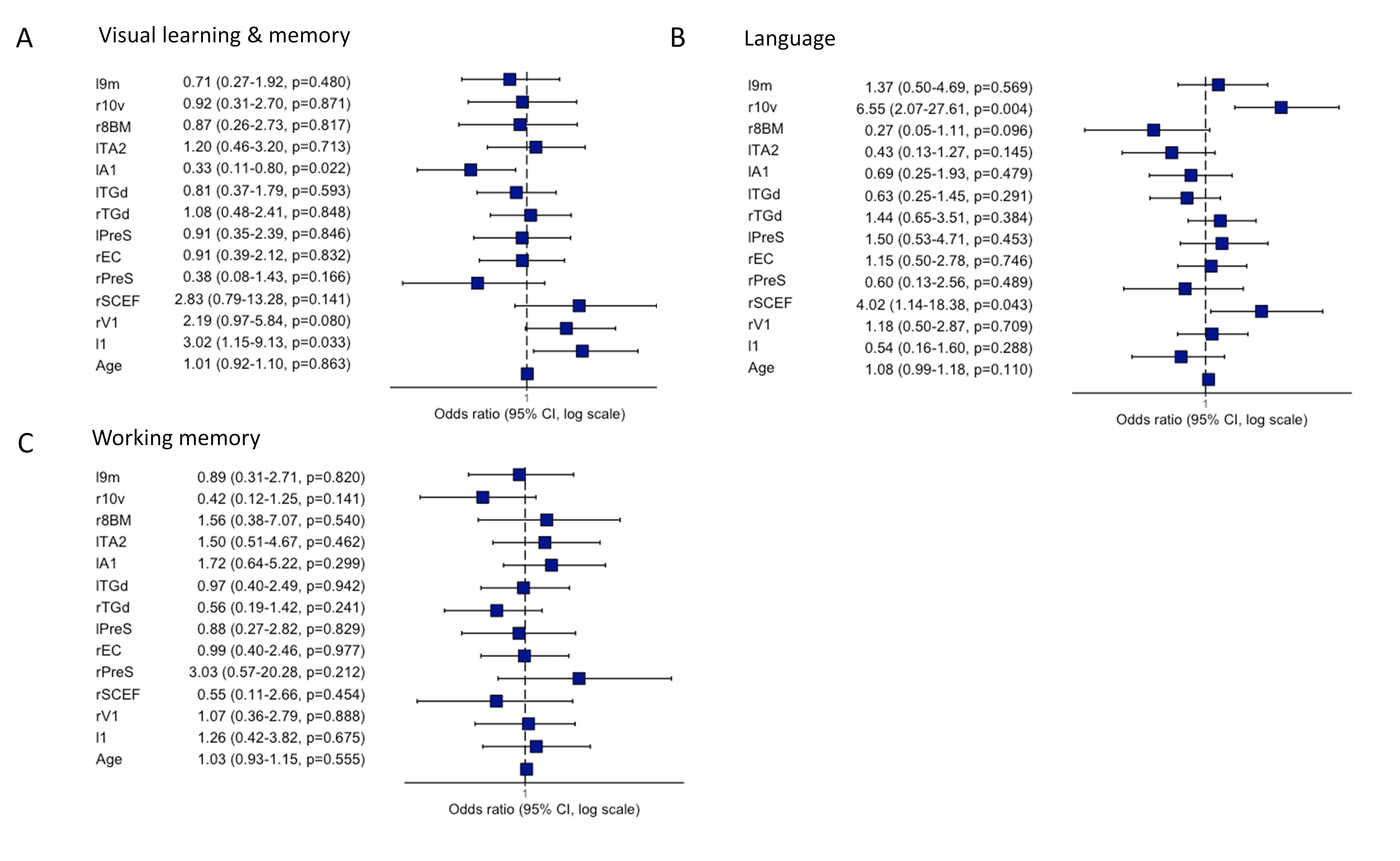

### Supplementary Figure 4

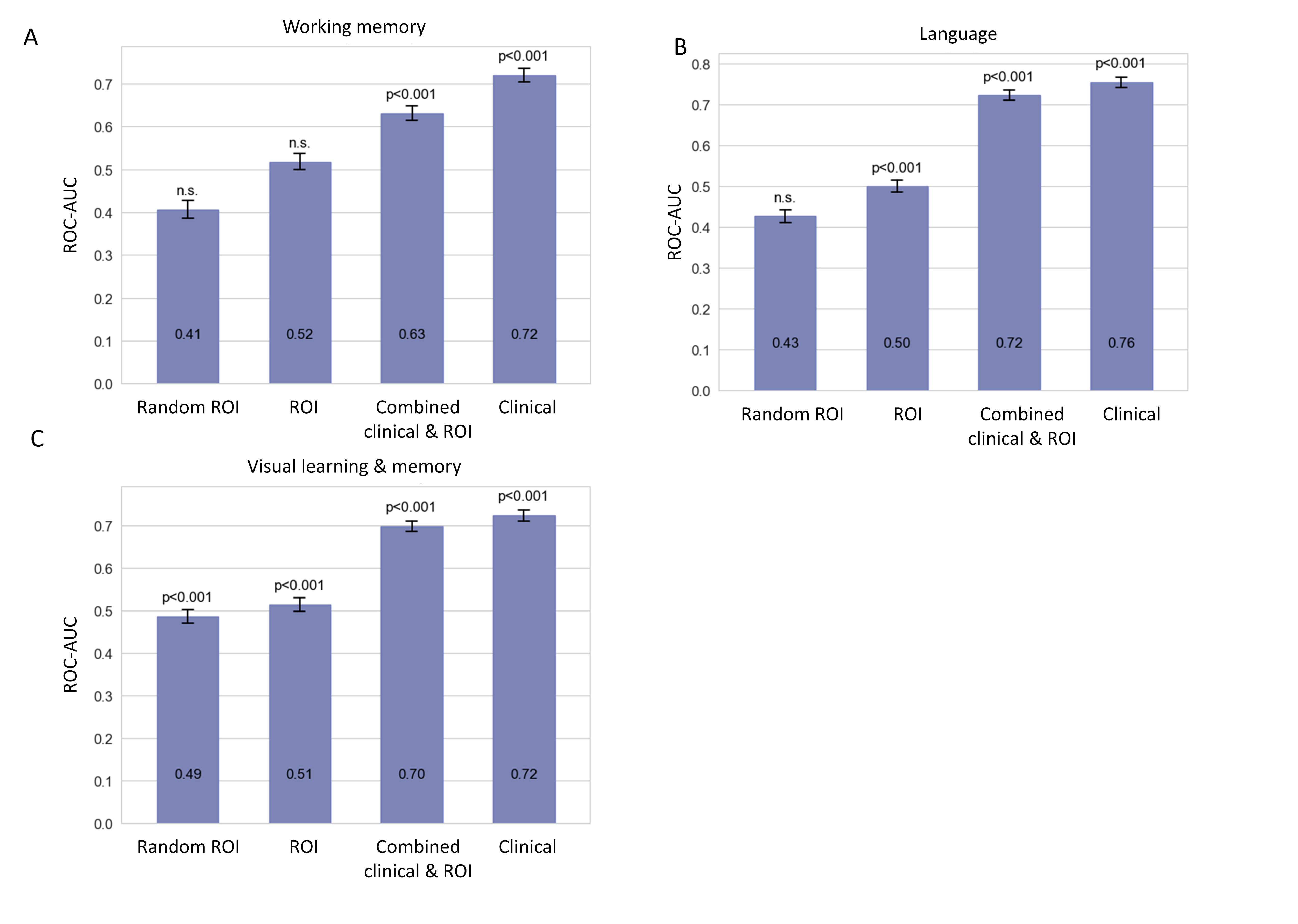

### Supplementary Figure 5

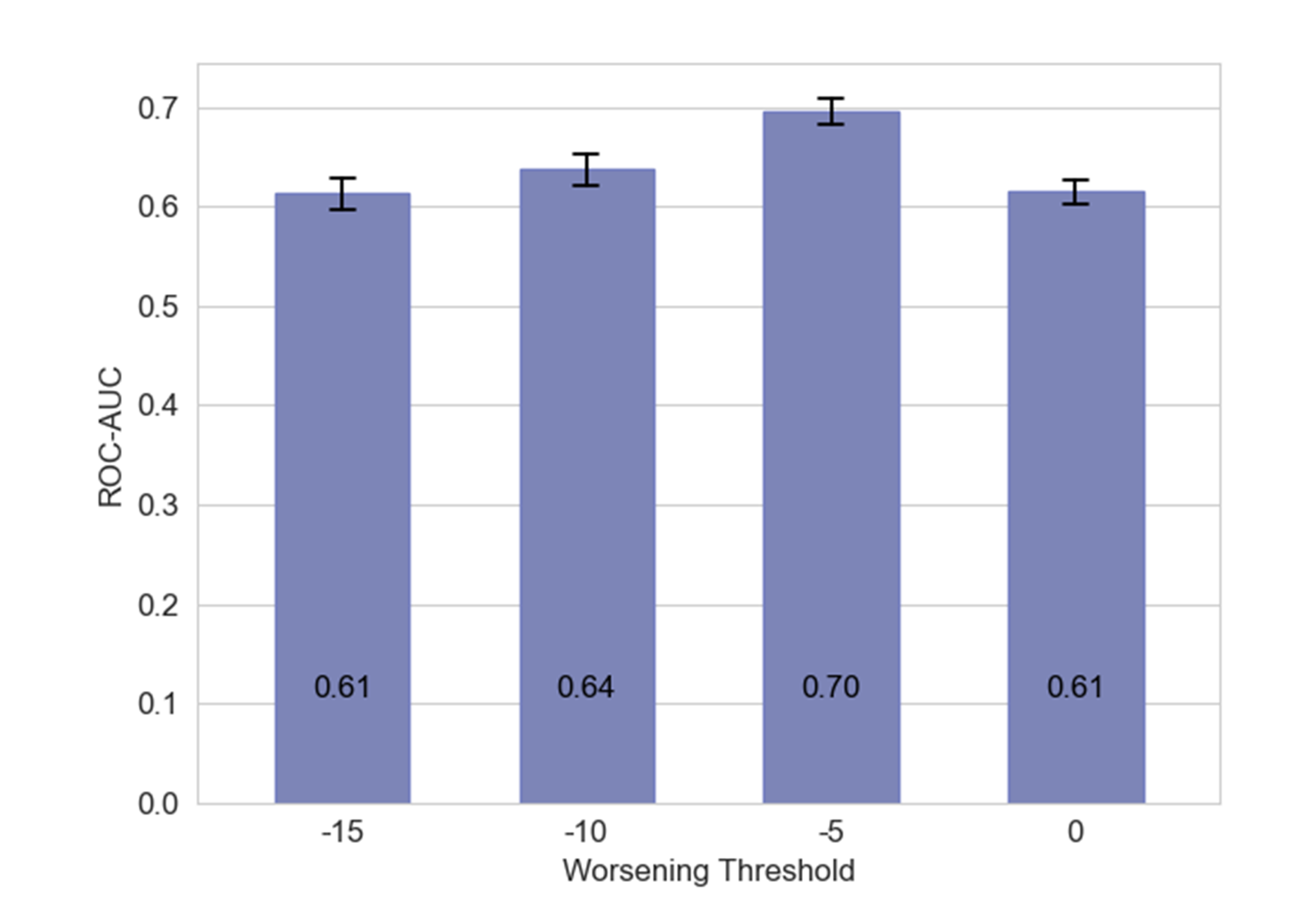
