## supplementary methods for "Cortical Thickness Patterns of Cognitive Impairment Phenotypes in Drug-Resistant Temporal Lobe Epilepsy"

**Supplementary material**

**Supplementary methods**

***Permutation testing procedure***

To assess the robustness of our results, we conducted a permutation test in addition to the standard ANOVA analysis. First, the cortical thickness labels within each patient were randomly shuffled (using NumPy). Second, the ANOVA analysis was run on the permuted data. These two steps were repeated a total of 100 times. Last, the empirical p-value was deemed significant if it fell into the 5^th^ percentile of p-value distribution in the permutation tests.

***Exploratory examination of post-surgical cognitive outcome prediction***

For patients who underwent epilepsy surgery, we conducted an additional exploratory analysis for 1-year post-surgical cognitive outcome prediction for each cognitive domain, except attention which was excluded due to missing data constraints. Cognitive worsening was defined as a decrease in post-surgical score of more than 5% compared to the pre-surgical score. For predictions, we utilized the combined ROIs (n=13) identified as most significantly connected to each cognitive phenotypes (distances calculated from cognitive phenotype to cortical GM regions that are below 1), as post-surgical change was similarly distributed across cognitive profiles and splitting into separate phenotypic groups would yield sample sizes that are insufficient for predictive modelling. First, descriptive LRMs were fitted to cognitive outcomes in each domain. Second, to determine if these regions along with clinical parameters could be predictive of cognitive changes, we applied 100 times repeated 5-fold nested cross-validation with recursive feature elimination and l2 regularizations (hyperparameters l2 regularization strength and number of features). Parameters used for clinically based prediction models included age, sex, disease duration, seizure onset side, presence of FBTCS, total number of ASMs, presurgical cognitive test percentile score, and presence of mesial temporal sclerosis on imaging.

To assess the performance of the models, we compared them to two random classifiers. The first established chance level by randomly assigning predictions classes to patients while maintaining the label prevalence of the data. The second classifier utilised random ROIs instead of the original factors selected as features (keeping n=13). This provided insight into whether the identified ROIs have improved predictive power compared general changes in brain cortical GM thicknesses in predicting post-surgical outcomes.

Different LRMs were compared using the receiver operating curve-area under the curve (ROC-AUC) values calculated from the hold-out dataset predictions of the cross-validation scheme. Non-parametric Mann-Whitney-U tests with Bonferroni correction for multiple comparison were used to evaluate differences between ROC-AUC from each model.

**Supplementary figures**

Supplementary figure 1: Effect sizes of cortical changes. The plots show thickness changes effect sizes of cortical grey matter regions that had significant differences from age- and sex-matched healthy controls for the three cognitive phenotypes. Negative change indicates cortical thinning compared to healthy controls, whereas positive change indicates increased cortical thickness compared to healthy controls.

Supplementary figure 2: Cortical thickness changes related to age. The plots show cortical grey matter regions with age related significant differences from age- and sex-matched healthy controls for the three cognitive phenotypes. Distance is defined as the average F-statistic / F-statistic, with shorter distances reflecting a more robust difference between cognitive impaired patient and healthy controls. Shorter distances are reflected by lighter shades of red.

Supplementary figure 3: Descriptive logistic regression models of cognitive post-surgical outcome for visual learning and memory, language, and working memory. Odds ratios for descriptive logistic regression classifying the change for each cognitive domain after surgery using the regions strongest associated with cognitive phenotype and age as independent variables. The cortical thicknesses are rescaled with respect to the mean and standard deviation of the healthy controls.

Supplementary figure 4: Predictive logistic regression models of cognitive post-surgical outcome for visual learning and memory, language, and working memory. Area under the receiver operating curve (ROC-AUC) of predictive logistic regression models with recursive feature reduction and l2 regularization for changes of verbal memory after surgery evaluated through repeated, nested cross-validation. Models displayed include: random ROI- predictor based on randomly chosen brain regions; ROI- predictor based on cognitive phenotype significantly associated regions; clinical- predictor based on clinical variables; combined clinical and ROI. P values indicate significant difference from a chance level classifier and from each other.

Supplementary figure 5: Area under the receiver operating curve (ROC-AUC) of region of interest (ROI) based logistic regression model predicting post-surgical verbal memory and learning outcome. Results applying different thresholds of post-surgical change are presented. P-value shows statistical significance from a chance level classifier.

**Supplementary table legends**

Supplementary table 1: type III ANOVA analysis used for directed network analysis. The From column contains the clinical origin node, the To column shows the region of interest independent variables, Weight is the resulting F score, and Bonferroni corrected, and non-adjusted p-values are presented. Table includes results for generalised, focal, and minimally impaired analyses.

Supplementary table 2: list of significantly affected regions in the cognitive impairment groups. Distances to cognitive phenotype, age, and MRI are given, as well as region of interest and brain area names.
